# Incidence and Risk Factors of Calcium Channel Blocker–Related Edema in Hypertensive Patients: A Multicenter Retrospective Cohort Study

**DOI:** 10.1101/2025.10.30.25339181

**Authors:** Koricho Simie Tolla, Gashaw Solela, Getachew Wondafrash, Abay Burusie, Gebi Agero, Dureti Desta Garoma, Wubshet Abraham Alemu, Bereket Sinshaw Engida, Surafel Mekasha Woldeyes, Berhanu Moges Abera, Mulualem Alemayehu Gebreselassie

## Abstract

**Background:** Hypertension is a major risk factor for cardiovascular disease and remains the leading cause of mortality worldwide. Calcium channel blockers (CCBs) are commonly used to lower blood pressure because they are effective and affordable. However, CCBs can cause vasodilatory adverse effects, including peripheral edema, which may lead to additional therapy and affect adherence. This study assessed the incidence and risk factors of CCB-related edema among hypertensive patients in Ethiopia.

**Methods:** This retrospective multicenter cohort study involved interviews and reviews of medical records of adults (aged ≥18 years) with essential hypertension who were prescribed calcium channel blockers (CCBs) between July 15 and August 14, 2025. A total of 292 participants were selected using systematic random sampling. A structured questionnaire was used to collect sociodemographic and clinical data. Descriptive statistics summarized baseline characteristics. Time-to-event analysis with the log-rank test assessed the duration from CCB initiation to edema onset. Binary and multivariate logistic regression analyses identified factors associated with CCB-related edema, and a *p*-value <0.05 was considered statistically significant.

**Results:** Among 292 participants (mean age 58.2 years; 53.4% female), 20.9% had diabetes mellitus and 16.8% had dyslipidemia. Amlodipine was the most frequently prescribed CCB (94.8%). Peripheral edema developed in 38.7% of patients, with a mean onset time of 8.3 weeks. In multivariate analysis, only longer daily standing duration was significantly associated with edema (AOR = 1.92; 95% confidence interval: 1.03–3.58; *p* = 0.041). Time-to-event analysis showed a progressive increase in edema risk with continued CCB use. Patients receiving amlodipine 10 mg daily had a greater (42.5% vs. 33%) and earlier risk of edema than those on 5 mg amlodipine daily; log-rank *p* = 0.003).

**Conclusions:** Peripheral edema occurred in over one-third of hypertensive patients on CCBs, appearing on average after eight weeks. Longer daily standing and higher CCB doses increased risk and earlier onset. Clinicians should inform patients and avoid unnecessary interventions. Further research on upright posture and edema is warranted.

## Introduction

Hypertension is defined as a persistent systolic blood pressure (SBP) of at least 130 mmHg or a diastolic blood pressure (DBP) of at least 80 mmHg and is associated with an increased risk of cardiovascular disease (CVD) events, including coronary heart disease, heart failure, stroke, and death [1]. The global prevalence of hypertension has doubled over the past three decades, with nearly two-thirds of affected individuals residing in low- and middle-income countries (LMICs) [2, 3]. Despite this high burden, treatment and control rates remain low in many regions, particularly in sub-Saharan Africa [2]. In Ethiopia, hypertension has become a growing public health problem, with urban prevalence among adults reported to be as high as 30.3% [4].

Randomized controlled trials (RCTs) have consistently demonstrated that antihypertensive therapy diminishes cardiovascular morbidity and mortality [5]. The Eighth Joint National Committee (JNC 8) guidelines recommend that initial therapy should include a thiazide diuretic, calcium channel blocker (CCB), angiotensin-converting enzyme (ACE) inhibitor, or angiotensin receptor blocker (ARB) [6]. Among these, CCBs and ARBs are the most frequently prescribed drug classes worldwide. Multiple studies have identified CCBs as the most commonly used antihypertensive medications [7]. Similarly, studies from Ethiopia have identified CCBs, along with ACE inhibitors and thiazide diuretics, as the most commonly prescribed antihypertensive agents [8, 9].

Dihydropyridine (DHP) CCBs are associated with vasodilatory adverse effects, including peripheral edema [10]. This edema is thought to result from increased capillary hydrostatic pressure due to greater dilation of pre-capillary arterioles compared to post-capillary venules [11]. Peripheral edema not only causes discomfort but may also lead to additional investigations, the prescription of loop diuretics, and poor adherence to therapy. Patients on DHP-CCBs are reported to be up to 60% more likely to receive loop diuretics than those on other antihypertensives [10, 12].

The reported incidence of CCB-induced peripheral edema varies widely. Meta-analyses of RCTs have shown a pooled incidence of around 10.7% [13], while some observational studies have reported rates as high as 29% [14, 15]. This variation may be due to differences in the design of the studies, the people who took part, or environmental or genetic factors. Several potential risk modifiers have been identified, including CCB type, dosage, duration of use, age, sex, comorbidities (such as diabetes, dyslipidemia, chronic obstructive pulmonary disease, cerebrovascular disease, and coronary artery disease), and upright posture [13, 16-18]. However, the influence of many of these factors remains inconsistent across studies.

Given the lack of local data and potential differences in patient characteristics, this study aimed to determine the incidence and identify risk factors of CCB-related peripheral edema among hypertensive patients treated at two teaching hospitals in Ethiopia. Findings from this study are expected to raise awareness among clinicians, improve patient counseling, and help reduce unnecessary investigations and additional drug therapy related to CCB-induced edema.

## Materials and methods

### Study design, area, and period

A multicenter retrospective cohort study was conducted by reviewing medical records and interviewing adult patients (aged ≥18 years) diagnosed with essential hypertension who were prescribed CCBs. The study was carried out from July 15 to August 14, 2025, at Asella Teaching and Referral Hospital (ATRH) and Yekatit 12 Hospital Medical College (Y12HMC). These hospitals were purposefully selected to capture diverse patient populations, with ATRH serving suburban and rural communities and Y12HMC serving an urban population. Both institutions have well-established hypertension follow-up clinics that provide continuous care, maintain comprehensive medical records, and are staffed by residents, internists, fellows, and subspecialists, ensuring reliable sources of clinical and patient information for this study.

### Source and study population

The source population included all adult patients (aged ≥18 years) diagnosed with essential hypertension and attending follow-up clinics at ATRH and Y12HMC who had been prescribed CCBs as part of their treatment regimen. The study population comprised those patients who attended follow-up during the study period and had complete medical records. Patients were excluded if they were non-compliant with treatment or if data on CCB usage or peripheral edema were missing.

### Eligibility criteria

Adult patients aged 18 years and above, diagnosed with essential hypertension and prescribed CCBs for at least six months, or who discontinued CCB therapy earlier due to the development of peripheral edema, were eligible for inclusion. All types of CCBs, including amlodipine and nifedipine, were considered. Patients with pre-existing conditions that could contribute to edema, such as heart failure, renal insufficiency, deep vein thrombosis, venous insufficiency, chronic liver disease, hypothyroidism, pregnancy, or other relevant conditions, were excluded. Additionally, patients with incomplete medical records regarding history, CCB use, or development of edema, as well as those unable to provide informed consent, were not included in the study.

### Sample size determination and sampling technique

The sample size was calculated based on a Nigerian prospective cohort study reporting a 29% incidence of peripheral edema among patients prescribed amlodipine or nifedipine [15]. Using a single population proportion formula with 95% confidence and 80% power, the initial sample size was 316. Considering that high-dose CCBs increase the risk of edema 2.8-fold [13], Fleiss’ formula with continuity correction yielded 318. With a total hypertensive patient population of approximately 2,500 at both centers, and 50% on CCB therapy, the source population was estimated at 1,250. Applying the finite population correction resulted in an adjusted sample size of 253. Accounting for a 10% non-response rate, the final sample size was 278.

A consecutive sampling technique was used, enrolling all eligible patients attending hypertension follow-up clinics during the study period until the target sample size was reached. Of the total sample, 52% (n = 145) were recruited from ATRH and 48% (n = 133) from Y12HMC. Systematic random sampling was used to select patients attending referral clinics during the study period. If the sampled patient was not a CCB user, the CCB user following him/her was selected as a sample.

### Study variables

The dependent variable was the incidence of calcium channel blocker (CCB)-related peripheral edema. The independent variables included sociodemographic factors (age, gender, and occupation), duration of daily upright posture, comorbid conditions (diabetes, chronic obstructive pulmonary disease, cerebrovascular accidents, coronary artery disease, dyslipidemia, and others), body mass index, and medication history. Medication history encompassed the type and dosage of CCBs prescribed, duration of therapy before discontinuation or edema development, and the use of other antihypertensive medications, including ACE inhibitors, angiotensin receptor blockers, thiazides, and beta-blockers. Among these, the specific dosage of CCB was considered the primary independent variable.

### Operational definitions

#### CCB user

An adult patient with essential hypertension who has used a CCB for at least six months or who discontinued CCB therapy due to the development of peripheral edema. The primary CCBs in this study were amlodipine and nifedipine.

#### Posture

The predominant body position maintained during daily activities, classified as upright, sitting, or recumbent/lying down.

#### Adherence to salt restriction

Consumption of ≤5 g of table salt per day (approximately one teaspoon), in line with World Health Organization recommendations.

### Data collection procedure

Data were collected using a structured questionnaire administered through the Kobo Toolbox platform. Demographic information and selected details on edema detection and progression were obtained via patient interviews, while comorbidities, physical examination findings, and medication histories were extracted from medical records. Data collection was performed by a team of four internal medicine residents and two internists, under the supervision of an internist, a cardiology fellow, and a nephrologist. Despite differences in training levels, the assessment of edema and completion of the questionnaire were considered straightforward, minimizing potential inter-observer variability.

### Data analysis

Data were checked for completeness and analyzed using SPSS version 27 (IBM Corporation, Armonk, NY, USA). Descriptive statistics, including frequency distributions and measures of central tendency and dispersion, were used to summarize demographic and clinical characteristics of the study population. Time-to-event (survival) analysis was performed to evaluate the interval from initiation of CCB therapy to the development of peripheral edema, stratified by CCB dose, with patients who did not develop edema during follow-up being censored. Binary logistic regression was used to estimate odds ratios for the occurrence of edema, while multivariable logistic regression adjusted for potential confounders and assessed the independent associations of multiple risk factors with the outcome. Statistical significance was defined as a p-value < 0.05.

### Data quality assurance

A pretest was conducted on 5% of the medical records to evaluate the clarity, feasibility, and consistency of the questionnaire. Data were collected by trained physicians under the supervision of the principal investigators. The completeness, consistency, and accuracy of the data were regularly monitored, and any discrepancies were promptly addressed to minimize information bias and ensure data reliability.

## Results

### Sociodemographic data

A total of 292 patients met the inclusion criteria and were included in the analysis. The mean age was 58.2 years, and 53.4% were female. Mean age was similar between participants who did and did not develop peripheral edema (58.27 vs. 58.22 years). The mean duration of hypertension was 47.9 months, ranging from 1 to 360 months. Regarding occupation, 37% of participants were retired, 20.5% were farmers, 14% were unemployed, and 10.3% were merchants (Table 1). On average, participants reported spending 2.9 hours per day standing (standard deviation [SD] = 2.0; range = 0–10 hours). The mean body weight was 68.0 kg (SD = 11.8; range = 40–115 kg).

**Table 1.** Socio-demographic characteristics and comorbidities of hypertensive patients taking CCB at ATRH and Y12HMC, Ethiopia, 2025 (N = 292)

| Variable | Number | Percentage |
| --- | --- | --- |
| Total patients (n) | 292 | 100.0 |
| Sex |  |  |
| Female | 156 | 53.4 |
| Male | 136 | 46.6 |
| Occupation |  |  |
| Retired | 108 | 37.0 |
| Farmer | 60 | 20.5 |
| Merchant/Trader | 30 | 10.3 |
| Office worker | 21 | 7.2 |
| Others | 25 | 8.6 |
| Comorbidities |  |  |
| Diabetes mellitus | 61 | 20.9 |
| Dyslipidemia | 49 | 16.8 |
| Stroke | 28 | 9.6 |
| Ischemic heart disease | 25 | 8.6 |
| Neuropathy | 24 | 8.2 |
| HIV Infection | 9 | 3.1 |
| Chronic kidney disease | 2 | 0.7 |
| Psychiatric condition | 2 | 0.7 |

Comorbidities were common, with diabetes mellitus in 20.9%, dyslipidemia in 16.8%, stroke in 9.6%, ischemic heart disease in 8.6%, and neuropathy in 8.2% of patients. Chronic kidney disease, HIV infection, and other conditions were less frequent, while 49.7% of participants had no additional medical conditions (Table 1). At the time of the most recent visit or edema assessment, blood pressure was controlled in 64% of participants, with a mean systolic BP of 134.6 mmHg (range 80–207) and a mean diastolic BP of 78.9 mmHg (range 51–130).

### Medication history and adverse events

The most commonly prescribed CCB was amlodipine (95%), followed by nifedipine (4.5%). High-dose CCB was administered to 57.2% of patients (Fig. 1). Concomitant antihypertensive therapy was used in 57.2% of participants, with ACE inhibitors prescribed to 49.1% and thiazide diuretics to 26.9% of these patients (Table 2).

**Table 2.** Additional antihypertensives used among hypertensive patients taking CCB at ATRH and Y12HMC, Ethiopia, 2025 (N = 292)

| **Concomitant antihypertensive drugs** | **Frequency** | **Percentage** |
| --- | --- | --- |
| ACEi | 82 | 49.1 |
| Thiazide diuretics | 45 | 26.9 |
| ACEi + thiazide diuretics | 20 | 12 |
| ARBs | 6 | 3.6 |
| ARBs + thiazide diuretics | 5 | 3 |
| ACEi + BB | 3 | 1.8 |
| ACEi + BB + others | 2 | 1.2 |
| Other combinations | 4 | 2.4 |
| Total | 167 | 100.0 |

**Abbreviations:** ACEi, angiotensin-converting enzyme inhibitor; ARB, angiotensin receptor blocker; BB, beta-blocker

**Fig. 1.**
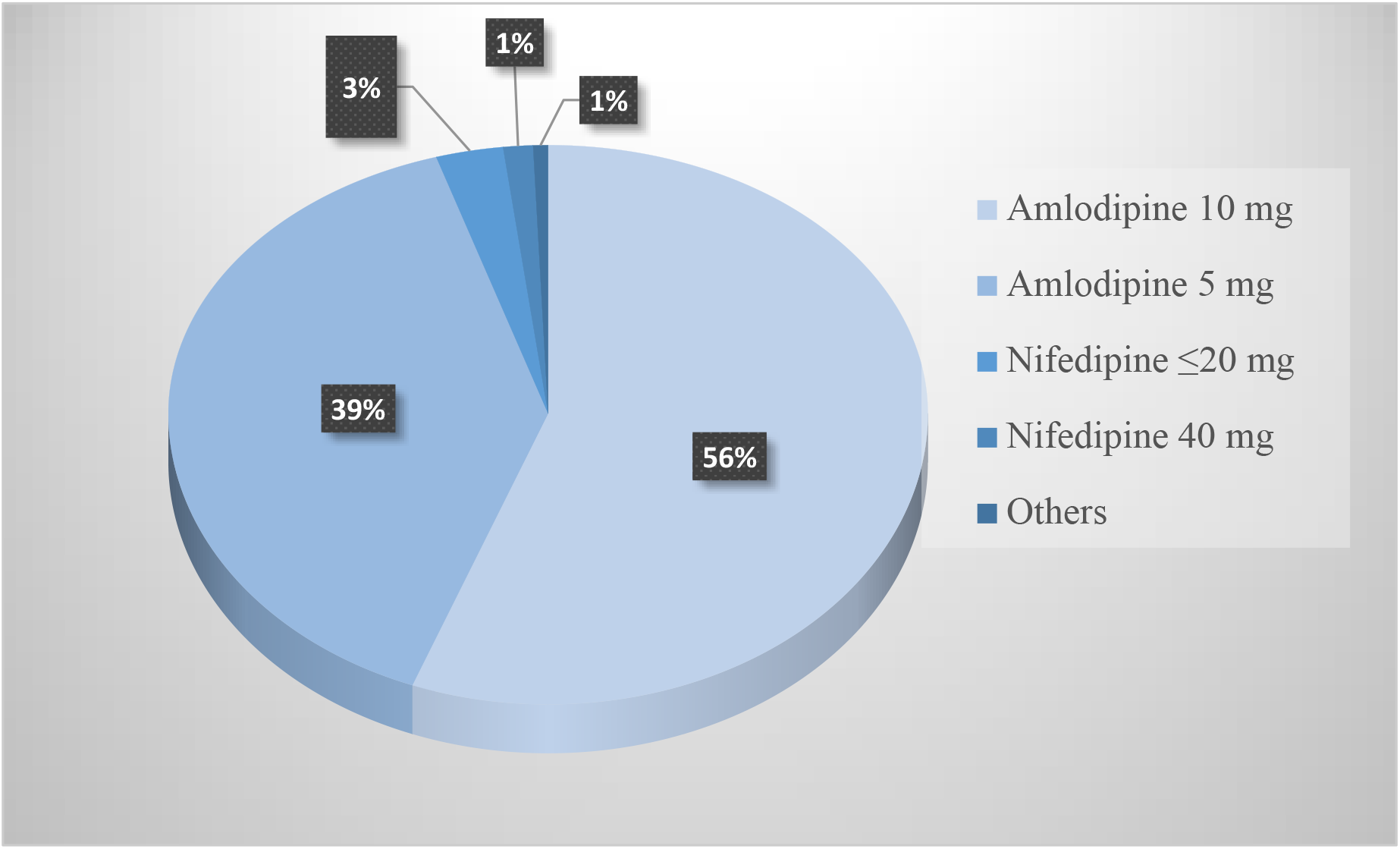
Pie chart for the types and daily doses of CCBs used among hypertensive patients at ATRH and Y12HMC, Ethiopia, 2025 (N = 292)

During follow-up, 38.7% of patients developed peripheral edema, with a mean onset of 8.3 weeks (range: 1–55 weeks) after initiating CCB therapy. Edema was predominantly bilateral and occurred more often in patients on high-dose therapy compared to low-dose therapy (42.5% vs. 33%). Patients self-reported edema in 84.9% of cases. Other commonly reported adverse effects included headache (14%), constipation (11.3%), and dizziness or lightheadedness (9.6%).

### Risk factors

Binary logistic regression showed that longer daily standing duration and high-dose CCB therapy were associated with edema development. In multivariate analysis, only daily standing duration remained significant (AOR: 1.92; 95% CI: 1.03–3.58; P = 0.041), while high-dose CCB approached significance (AOR: 1.58; 95% CI: 0.97–2.57; P = 0.068). No comorbid conditions or other adverse effects, including headache and constipation, were associated with edema (Table 3).

**Table 3.** Logistic regression on risk factors for edema among hypertensive patients taking CCB at ATRH and Y12HMC, Ethiopia, 2025 (N = 292)

| Variable | COR (95% CI) | p-value | AOR (95% CI) | p-value |
| --- | --- | --- | --- | --- |
| Age | 1.01 (0.99-1.03) | 0.48 | - | - |
| Sex (male) | 1.60 (0.92-2.80) | 0.099 | - | - |
| Duration of daily standing | 1.2(1.02-1.3) | 0.02 | 1.92(1.026-3.58) | 0.041 |
| High-dose CCB | 1.45 (1.018-2.06) | 0.039 | 1.58(0.97-2.57) | 0.068 |
| Use of other antihypertensive medications | 0.60 (0.36-1.02) | 0.06 | - | - |
| Controlled BP | 0.72 (0.44-1.17) | 0.18 | - | - |
| Other CCB side effects | 1.93 (0.67-5.56) | 0.23 | - | - |
| Physical exercise | 1.66 (0.55-4.99) | 0.37 | - | - |
| Adherence to salt restriction | 1.37 (1.56-3.35) | 0.49 | - | - |
| Current smoking status | 0.67 (.062-7.18) | 0.74 | - | - |
| Daily alcohol consumption | 4.91 (0.74-32.66) | 0.10 | - | - |

**Abbreviations:** BP, blood pressure; CCB, calcium channel blocker

### Management of CCB-related edema

Following edema onset, management strategies varied. Drug adjustment, including discontinuation, was performed in 58.4% of patients, and additional investigations were conducted in 23.9%. Diuretics were added in 22.3% of cases, whereas no patients received ACE inhibitors or ARBs. A minority of patients (17.7%) continued therapy with conservative measures, such as lifestyle modifications.

### Time-to-event analysis

Time-to-event analysis, stratified by daily CCB dose (high vs. low), showed a progressive increase in the risk of edema with longer duration of CCB use. Participants receiving amlodipine 10 mg experienced a higher and earlier risk of edema compared to those taking 5 mg (Fig. 2). The difference between the groups was statistically significant by the log-rank test (P = 0.003; Log Rank [Mantel-Cox] = 15.703). Analysis was restricted to participants taking amlodipine, which accounted for nearly 95% of the study population.

**Fig. 2.**
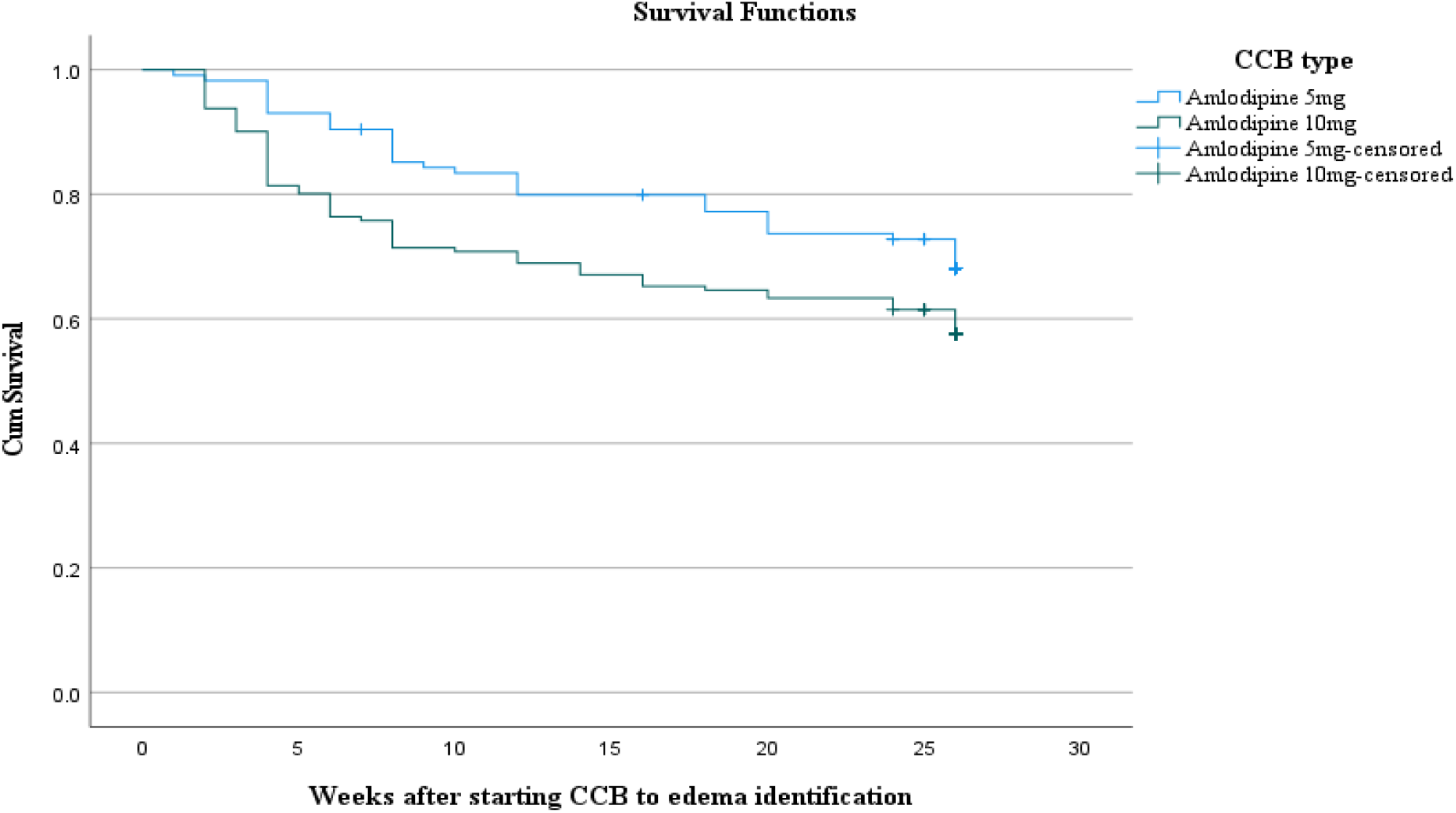
Kaplan-Meier curve for development of edema among hypertensive patients taking CCB at ATRH and Y12HMC, Ethiopia, 2025 (N = 292)

## Discussion

In this study, we retrospectively examined the incidence of CCB-related edema among 292 patients with hypertension at two hospitals in Ethiopia. The mean age of 58.2 years was comparable to that reported in one of the large meta-analyses by Makani et al. [13]. Based on the results of this study, the incidence of CCB edema was more common among the Ethiopian population than reports from other locations. Incidence rates from studies and meta-analyses ranged from 10.7% to 29%, which are lower compared to our finding of 38.7% [13-15].

The average time to identify edema in our study was 8.3 weeks (58.1 days), which is closer to the Indian study of 69 days [14] compared to a much shorter duration, such as 27.3 days, as reported by a different study [20]. This difference may be explained by the delayed reporting by patients, who were responsible for identifying edema in 85% of cases. This is supported by the fact that edema was reported as early as 1 week after initiating CCB in this study. This suggests that active monitoring for edema by physicians after initiating CCBs could lead to earlier detection.

In agreement with several other studies, this study also showed significantly higher incidence of edema in patients using higher doses of CCB, particularly in participants taking amlodipine 10 mg compared to 5 mg [13]. There is also data that supports a progressive increase in the incidence of edema with longer duration of CCB use, which is the case in our study, as illustrated by the Kaplan-Meier curve in the results section [15]. Not just higher incidence, this curve also showed earlier onset of edema among high-dose CCB users. This is supported by the biological plausibility of high-dose CCB resulting in a higher effect on precapillary vessels and subsequently increasing capillary hydrostatic pressure and edema.

Several modifiers of CCB-related edema have been well reported by studies, including upright posture, warmth, older age, and female gender [21]. Our study showed a significant association with the duration of upright posture. This is due to the upright posture providing an additional gravitational contribution to increased hydrostatic pressure and the development of edema [16, 17]. One study showed an association of CCB edema with comorbid conditions, including diabetes, COPD, coronary artery disease, and dyslipidemia, which our study didn’t evidence. This is likely due to the fact that, except for diabetes, other comorbid conditions were less frequent in our study.

The use of RAS inhibitors, such as ACE inhibitors, has been shown to reduce the incidence and severity of CCB-related edema [13, 16, 17]. Although there was a tendency to decrease the risk of CCB edema with concomitant use of ACEi and/or thiazides with an odds ratio of 0.60 (95% 0.36-1.02, p = 0.06), it didn’t reach statistical significance. This may be due to the low number of participants using these medications.

Lack of acknowledgment of edema as an adverse effect of CCBs by clinicians might result in inadvertent use of additional interventions. One of the adverse effects of CCB-related edema is the sequential use of diuretics, which may expose patients to drug-related side effects like acute kidney injury. A large cohort study reported that CCB users experienced 60% higher rates of being subsequently dispensed a loop diuretic compared to other antihypertensive drug users [12]. Although we cannot directly compare this figure, our study found that 22.3% of participants were prescribed to use diuretics. Additionally, nearly 24% of our patients underwent additional laboratory and imaging tests after the detection of edema.

## Conclusion

CCB-related edema is common in Ethiopian patients with hypertension and is associated with longer daily standing and higher doses. Many patients undergo unnecessary tests or receive diuretics for this side effect. Clinicians should recognize its high incidence, inform patients, and avoid unnecessary interventions. Further research on the role of daily upright posture in edema development is warranted.

## Ethics approval and consent to participate

Ethical clearance was obtained from the College of Health Science, Arsi University, Ethical Review Committee (Ref. No: A/U/H/S/C/120/13619/17). Written informed consent was obtained from all participants. Data were anonymized to protect confidentiality, and only relevant research information was collected. Clinical care and research activities were strictly separated, with treating physicians not involved in data collection or analysis. No therapeutic interventions were performed. As this was a non-interventional study, a clinical trial number was not required. The study adhered to the Declaration of Helsinki.

## Consent for publication

Not applicable.

## Data availability statement

The datasets used or analyzed during the current study are available from the corresponding author upon reasonable request.

## Author contributions

KST was involved in conceptualization, supervision, methodology, formal analysis, and writing – original draft. GS, GW, AB, and GA were involved in supervision, methodology, formal analysis, and writing – review and editing. DDG, WAA, BSE, SMW, BMA, and MAG were involved in formal analysis, validation, and writing – review and editing. All authors reviewed and approved the final manuscript.

## Acknowledgments

The authors extend their gratitude to the study participants and the healthcare personnel at ATRH and Y12HMC referral clinics for their valuable support.

